# Epidemiology of seasonal coronaviruses: Establishing the context for COVID-19 emergence

**DOI:** 10.1101/2020.03.18.20037101

**Authors:** Sema Nickbakhsh, Antonia Ho, Diogo F.P. Marques, Jim McMenamin, Rory N. Gunson, Pablo R. Murcia

## Abstract

Public health preparedness for coronavirus disease 2019 (COVID-19) is challenging in the absence of setting-specific epidemiological data. Here we describe the epidemiology of seasonal human coronaviruses (sCoVs) and other cocirculating viruses in the West of Scotland, UK. We analyzed routine diagnostic data for >70,000 episodes of respiratory illness tested molecularly for multiple respiratory viruses between 2005 and 2017. Statistical associations with patient age and sex differed between CoV-229E, CoV-OC43 and CoV-NL63. Furthermore, the timing and magnitude of sCoV outbreaks did not occur concurrently and coinfections were not reported. With respect to other cocirculating respiratory viruses, we found evidence of positive, rather than negative, interactions with sCoVs. These findings highlight the importance of considering cocirculating viruses in the differential diagnosis of COVID-19. Further work is needed to establish the occurrence/degree of cross-protective immunity conferred across sCoVs and with COVID-19, as well as the role of viral coinfection in COVID-19 disease severity.

## Introduction

In March 2020, the WHO declared the global spread of coronavirus disease 2019 (COVID-19), caused by a human coronavirus (SARS-CoV-2) which emerged in China in December 2019, a pandemic [1]. Predicting the public health impact of pathogens with recently acquired human-to-human transmissibility is a challenge. Currently, the fate of COVID-19 remains unclear; understanding the likely age and seasonal profiles of infection risks will be critical to inform effective surveillance and control strategies.

During the early phase of an outbreak, in the absence of detailed country-specific knowledge, preliminary risk estimates may be gauged from endemic pathogens with similar modes of transmission. The infection incidence and levels of severe illness associated with COVID-19 remains unclear. In this instance, epidemiological data on seasonal human coronaviruses (sCoVs) may provide valuable information about individuals and seasonal conditions favored by an invading coronavirus.

To date, emergent zoonotic human coronaviruses associated with high case-fatality ratios have not achieved persistence in the human population. Severe acute respiratory syndrome coronavirus (SARS-CoV) emerged in 2002 and spread rapidly around the globe before being successfully contained in 2003 [2]. Conversely, Middle East respiratory syndrome coronavirus (MERS-CoV) has continued to cause sporadic cases predominantly in healthcare settings since its discovery in 2012, but has not demonstrated sustained community transmission [3]. In contrast, CoV-229E, CoV-NL63, CoV-OC43 and CoV-HKU1 are common cocirculating sCoVs predominantly associated with mild infection of the upper respiratory tract [4].

A key determinant governing the invasion and persistence success of a new pathogen is the abundance of susceptible hosts. Such population susceptibility may be difficult to define due to pre-existing cross-protective immunity in individuals previously exposed to antigenically related pathogens, as demonstrated for pandemic influenza A H1N1 in 2009 [5]. Furthermore, the potential for heterologous interactions among taxonomically broad groups of respiratory viruses is also recognized [6–11]. A good epidemiological understanding of cocirculating viruses will provide valuable information on the potential for immune, or otherwise mediated, virus-virus interactions and consequences for population susceptibility.

To date, epidemiological knowledge surrounding sCoVs has been limited for many settings owing to their historic association with mild illness. However, some laboratories have adopted sCoV testing as part of routine multiplex diagnostic screens [12–15], following an increased recognition of the associated disease spectrum. We previously reported on the comparative epidemiologies of acute viral respiratory infections, and the potential for virus-virus interactions, based on multiplex RT-PCR testing in the West of Scotland [6, 16]. Here we provide further detail on sCoVs differentiated at the species level (sCoV types) over an extended timeframe and discuss key potential implications for COVID-19 virus emergence in Scotland, United Kingdom (UK).

## Methods

### The study population

Routine molecular testing for CoV-229E, CoV-OC43 and CoV-NL63 using multiplex real-time RT-PCR methods was conducted between 1^st^ January 2005 and 30^th^ September 2017 by the West of Scotland Specialist Virology Centre (WoSSVC) in NHS Greater Glasgow and Clyde, the largest Scottish Health Board serving a population of ∼1.2 million [17]. The respiratory virus screen also simultaneously detected influenza A virus, influenza B virus, respiratory syncytial virus, human adenoviruses, human rhinoviruses, human metapneumovirus, and parainfluenza virus types 1-4. The CoV-HKU1 assay was discontinued in 2012 due to low levels of detection. Most clinical specimens (91%) were obtained from the upper respiratory tract (the majority being nasal and/or throat swabs).

During the study period, 107,174 clinical respiratory samples, from 64,948 individual patients, were received by WoSSVC for testing. For patients with two or more samples submitted (24.5% of patients), the PCR test data were aggregated into individual episodes defined as a 30-day period from the collection date of the first sample. This generated 84,957 episodes of respiratory illness for analysis. Most episodes, 93% out with the three major waves of pandemic influenza A(H1N1)pdm09 virus circulation in the UK (summer 2009, influenza seasons of 2009/10 and 2010/11), were tested for all eleven groups of respiratory virus. Of 84,957 episodes of respiratory illness, 10,438 were not tested for coronavirus (98% during the three major waves of pandemic influenza) and thus were excluded from analyses centered on sCoVs [18]. Among the remaining 74,519 episodes of illness, a further 278 were either tested for CoV-HKU1 or the coronavirus was untyped; these episodes were excluded from analyses differentiating sCoV type. See Figure 1 for a summary of the data subsets.

**Figure 1:**
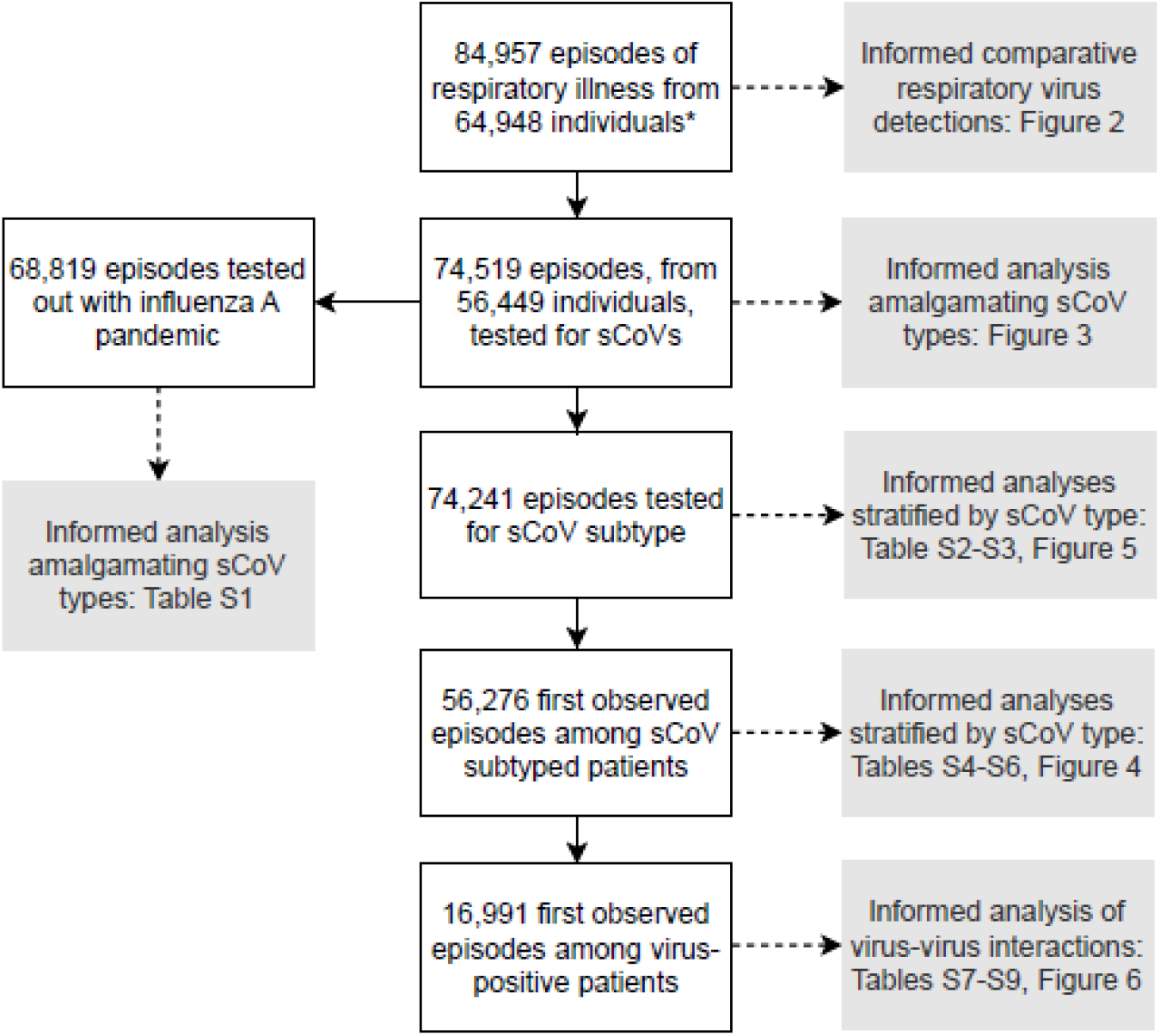
Data flow diagram summarising patient subsets informing each analysis. *Patients molecularly tested for respiratory viruses using real-time multiplex RT-PCR in NHS Greater Glasgow and Clyde, Scotland, UK between 1^st^ January 2005 and 30^th^ September 2017.

### Statistical modelling analyses

Of 74,241 patient episodes of respiratory illness with sCoV subtyping, 8,912 patients experienced multiple episodes over the study timeframe. In such cases, we retained the first observed episode to remove patient-level clustering, leaving 56,276 patient observations for analysis (Figure 1). We used multivariable logistic regression to investigate associations between each sCoV type (CoV-229E, CoV-OC43 and CoV-NL63) and patient age (categorical), sex (binary), healthcare service (binary), time period with respect to the three major waves of pandemic influenza in the UK (categorical; pre-pandemic: January 2005 – April 2009, pandemic: May 2009 – February 2011, post-pandemic: March 2011 – September 2017) and season (categorical). Statistical interactions between patient covariates and healthcare service were assessed. An alpha level of 5% was used to determine statistical significance of all model coefficients. The fitted models, incorporating age-healthcare service interactions were used to generate average predicted probabilities of virus detection by age and healthcare setting.

In addition, we used multivariable logistic regression to investigate interactions between each sCoV and other groups of respiratory viruses at the within-host scale. These analyses were based on 16,991 virus-positive episodes of respiratory illness, retaining the first observed episode of illness for patients with multiple episodes. Virus-negative patients were excluded to eliminate the influence of Berkson’s bias, which may lead to spurious inference of disease-disease associations when estimated from routine healthcare data [19]. Specifically, these analyses tested whether the odds of a given virus (‘exposed’) coinfecting with a given sCoV differed from the average odds among the remaining groups of viruses (‘non-exposed’), thereby assessing non-random mixing among the virus population.

Three models were fitted, one for each of CoV-229E, CoV-OC43 and CoV-NL63 (response variables). The analyses adjusted for patient age, sex, primary or secondary/tertiary healthcare service, time period with respect to pandemic influenza (as above), and the monthly background prevalence of the sCoV (response variable) to eliminate spurious virus-virus associations owing to unrelated sources of seasonality. Holm’s method was used to correct p-values for multiple comparisons (ten virus-virus interaction hypotheses per model).

All analyses were conducted in R software version 3.4.4 [20]. Logistic regression modelling was conducted using the ‘glm’ function and predicted probabilities were computed using ‘ggaverage’ from the ‘ggeffects’ package [21].

## Results

### How common are seasonal coronaviruses among people with respiratory illness?

Among 84,957 episodes of respiratory illness, 79.0% were sampled at secondary/tertiary healthcare services (hospital inpatients and outpatients), and 21.0% from primary healthcare services (general practice [GP]). The sex distribution was approximately equal, with 51.6% of patients’ female, and the median age was 33.1y (IQR; 1^st^ quartile - 3^rd^ quartile=5.6y-59.1y).

Seasonal coronaviruses (sCoVs) were detected in 4.0% (2,958/74,519) of tested patients overall, contributing to 10.7% (2,958/27,734) of all respiratory virus detections. Figure 2 summarizes the contribution of sCoVs to the total viral detections in the patient population during each influenza season (October – May) from 2005 to 2016. The most common virus detections overall among virus-positive patients were influenza (range: 13.4% to 34.0%, excluding pandemic influenza waves of 2009/10 and 2010/11) and RSV (range: 10.1% to 21.9%) followed by sCoVs (range: 7.7% to 17.4%) (Figure 2).

**Figure 2:**
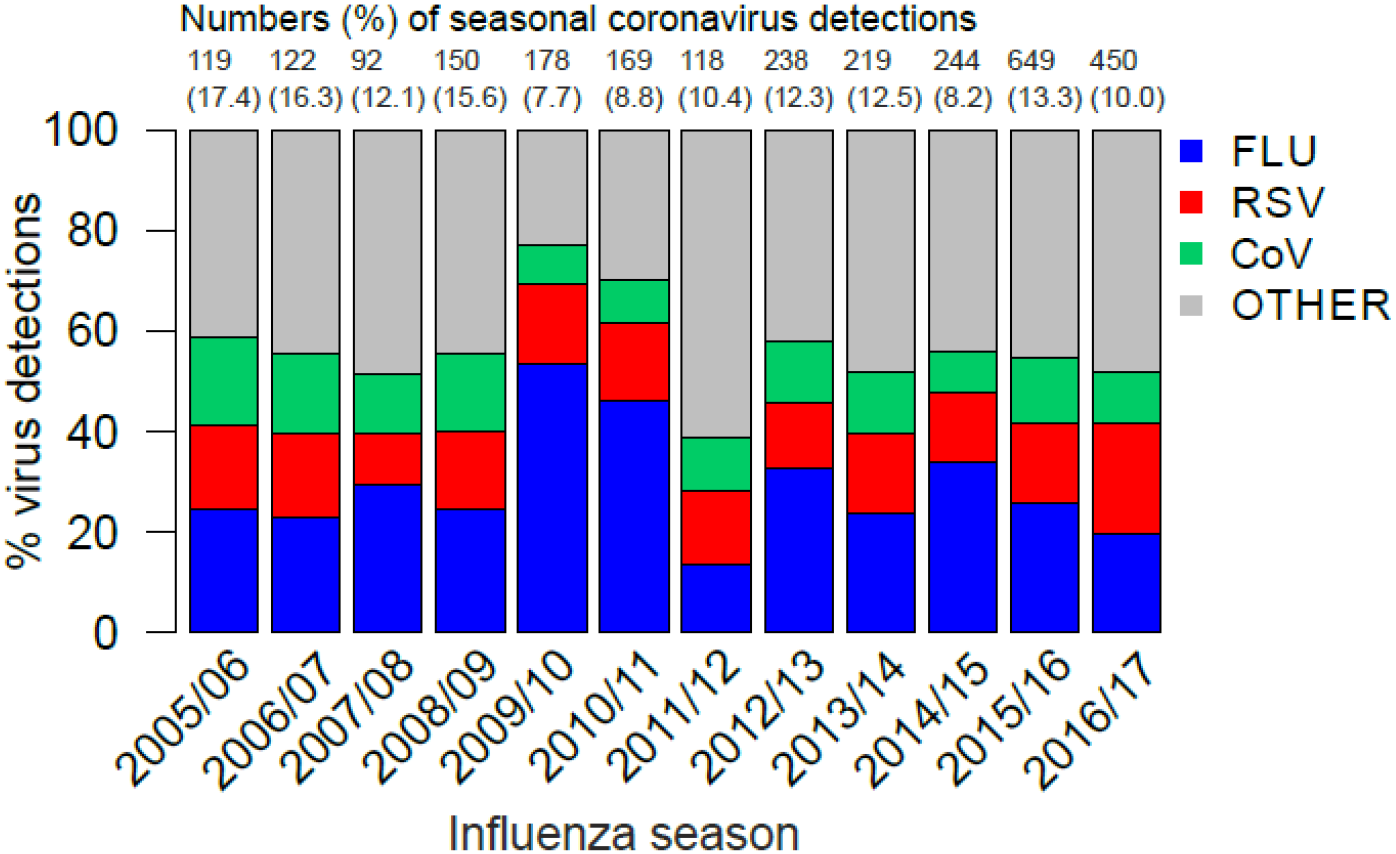
Percentage of viral respiratory infections attributed to human coronaviruses and other common respiratory viruses during each influenza season (October – May) from 2005/06 until 2016/17 based on 84,957 episodes of respiratory illness. FLU=influenza A and influenza B viruses combined; RSV=respiratory syncytial virus; CoV=human coronaviruses (CoV-229E, CoV-OC43, CoV-NL63 and CoV-HKU1 combined); OTHER=human adenoviruses, human rhinoviruses, human metapneumovirus, or parainfluenza viruses type 1-4. Note: years of major pandemic influenza A H1N1 virus circulation (2009/10 and 2010/11) must be viewed with caution due to high levels of partial testing. Testing for CoV-HKU1 was discontinued in 2012.

Numbers of sCoV detections increased post pandemic influenza (March 2011 – September 2017) likely owing to enhanced virological testing of acute respiratory illnesses; overall numbers of sCoV detections rose from 545 pre-pandemic to 2072 post-pandemic. However, a decrease in prevalence among the tested population was observed, from 4.27% to 3.70%, and with varying patterns at the individual sCoV level (supplementary Table S1). The most prevalent detection was CoV-OC43, both pre- and post-pandemic (supplementary Table S1). CoV-HKU1 was present at a very low prevalence of 0.3% overall (124 out of 36,652 episodes tested until the assay was discontinued in 2012) and was therefore excluded from further analyses.

### In whom are seasonal coronaviruses most frequently detected?

Despite a greater number of sCoV detections among patients admitted to hospital, the prevalence among those tested was greater among patients attending GP’s (5.3%; 673/12,670) than hospitals (3.7%; 2,285/61,849). Figure 3 summarizes age distributions. Cases of sCoV in children under five and the elderly were disproportionately represented among patients admitted to hospital, compared to a more uniform distribution among GP attendees, closely following the overall tested population (Figure 3A). A different sex bias was found among adults depending on the healthcare setting; more female cases (primary care) versus males (secondary/tertiary care) (Figure 3B). This pattern was consistent when comparing % detections among positive patients across sCoV types: 59.2% (CoV-229E), 55.6% (CoV-OC43) and 59.8% (CoV-NL63) of cases detected in primary care were female, whilst 54.7% (CoV-229E), 51.1% (CoV-OC43) and 56.7% (CoV-NL63) cases detected in secondary/tertiary care were male (supplementary Table S2).

**Figure 3:**
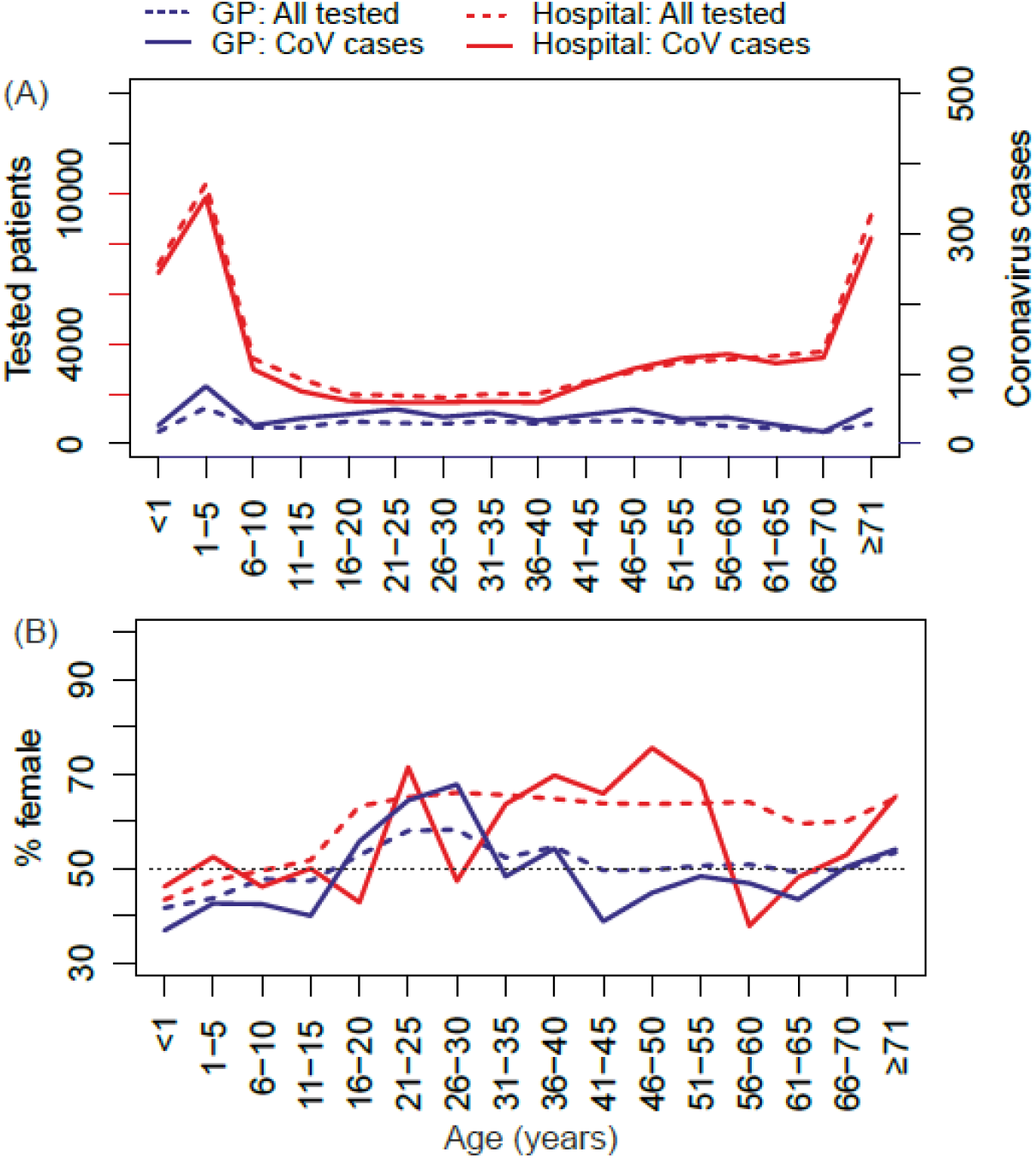
Age distributions of human coronavirus cases. (A) comparing numbers positive and tested among patients attending GP’s (primary care) and hospitals (secondary/tertiary care), and (B) percentage of female patients. Note the different y-axis scale for coronavirus cases in figure A. GP=General Practice surgery; Hospital=inpatients and outpatients.

The median age varied from 20.9y (IQR=2.7y-50.2y) for CoV-NL63, to 39.9y (IQR=5.0y-62.5y) and 43.3y (IQR=16.5y-60.4y) for CoV-OC43 and CoV-229E, respectively. The age-specific prevalences of sCoVs among the tested population are summarized in supplementary Tables S3-S4; a greater variation across ages was found for CoV-229E (Coefficient of Variation [CV]=40.4%) and CoV-NL63 (CV=33.8%) compared to CoV-OC43 (CV=13.6%) for primary care, with less variation for secondary/tertiary care (CV=29.96% for CoV-229E and CV=28.0% for CoV-NL63, compared to CV=17.10% for CoV-OC43).

Statistical modelling analyses further confirmed differences in age and sex associations according to sCoV type, and a greater chance of sCoV detection among GP attendees than patients admitted to hospital (supplementary Tables S5-S7). No evidence of significant effect modification between patient age or sex and healthcare service setting was found (statistical interaction terms p>0.05; results not shown). Figure 4 summarizes average age-specific predicted probabilities with statistical interactions incorporated. In summary, we observed a trend towards increasing probability of CoV-229E with age (Figure 4A), greater probabilities of CoV-OC43 at the extremities of age (Figure 4B), and decreasing probability of CoV-NL63 with age (Figure 4C). These age patterns were broadly consistent across patient sex and healthcare settings; although we note that 95% confidence intervals overlapped across all ages except for the hospital setting. A borderline significant sex effect was found for CoV-229E, with detections more likely among males (supplementary Table S5).

**Figure 4:**
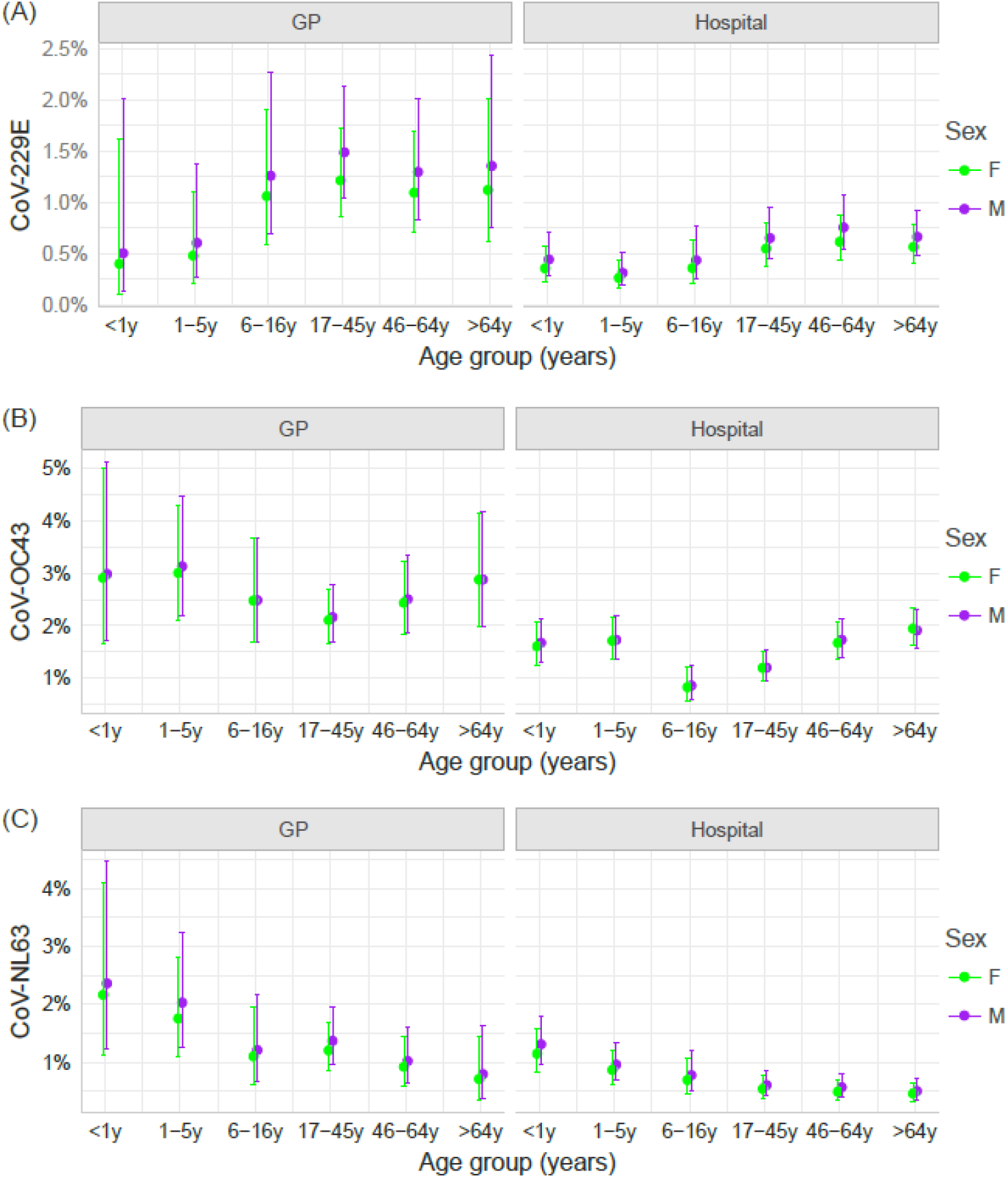
Average age-specific predicted probabilities (pp) of human coronavirus (CoV) detections by patient sex and healthcare service setting. GP=General Practice (primary care); Hospital=inpatients and outpatients (secondary/tertiary care); F=female; M=male. Derived from multivariable logistic regression models incorporating statistical interaction between patient age and healthcare service (see supplementary Tables S5-S7 for model results without statistical interactions).

### Do seasonal coronaviruses exhibit similar seasonalities?

Figure 5 shows the monthly prevalences of sCoVs detected among the patient population. These are winter pathogens in the UK, on average peaking between January and March. However, there were notable variations between sCoV types and between years. Overall, CoV-OC43 was the most prevalent detection among the tested population in each influenza season. Differences were also observed in periodicities; prior to the first wave of pandemic influenza in 2009, CoV-229E peaked biennially, but subsequently exhibited longer inter-peak periods, particularly between 2013 and 2016.

**Figure 5:**
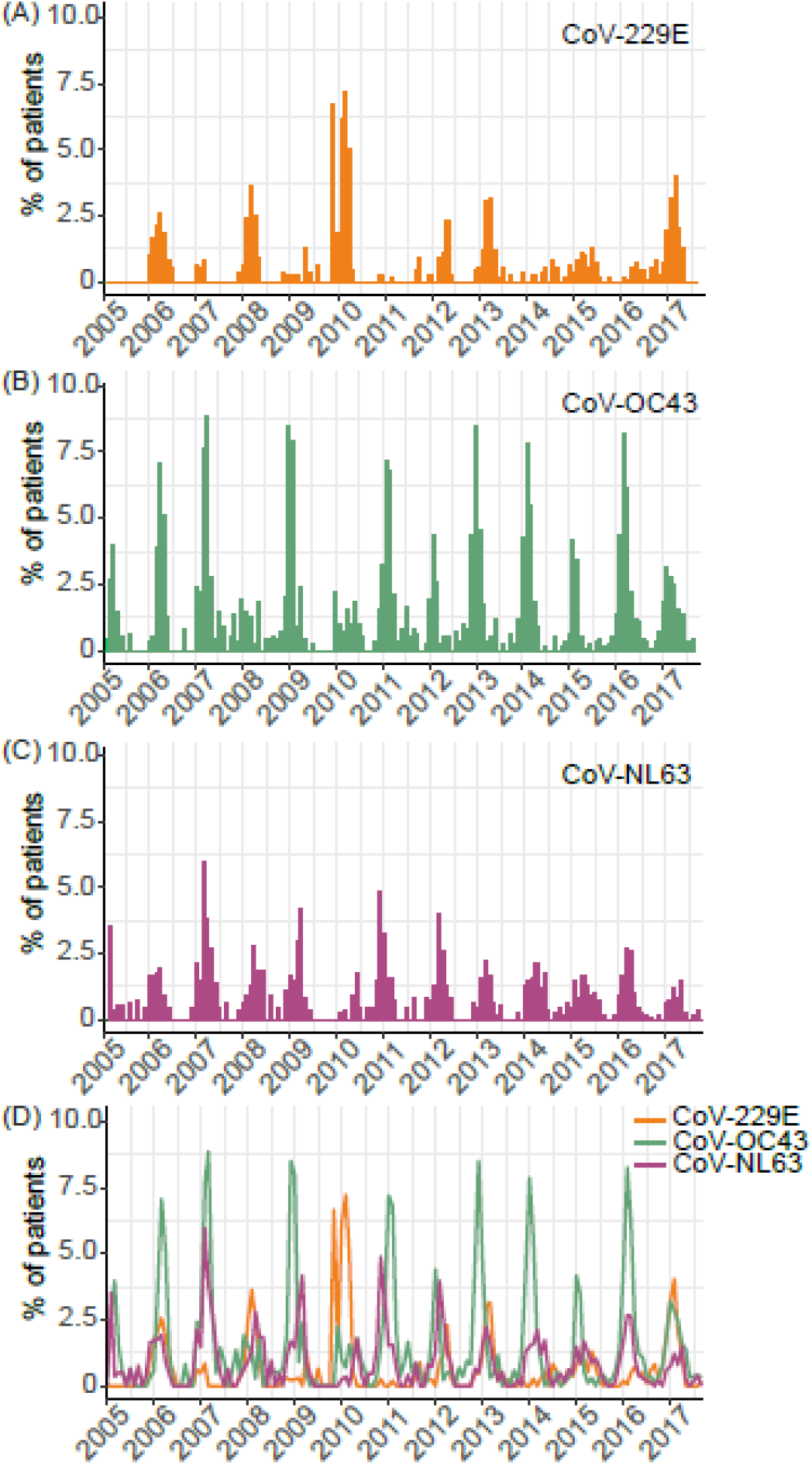
Monthly prevalence (%) of seasonal coronaviruses (sCoVs) detected among patients with respiratory illness virologically tested in NHS Greater Glasgow & Clyde, Scotland, UK, between January 2005 and September 2017: (A) CoV-229E, (B) CoV-OC43, (C) CoV-NL63, and (D) comparing all sCoVs.

**Figure 6:**
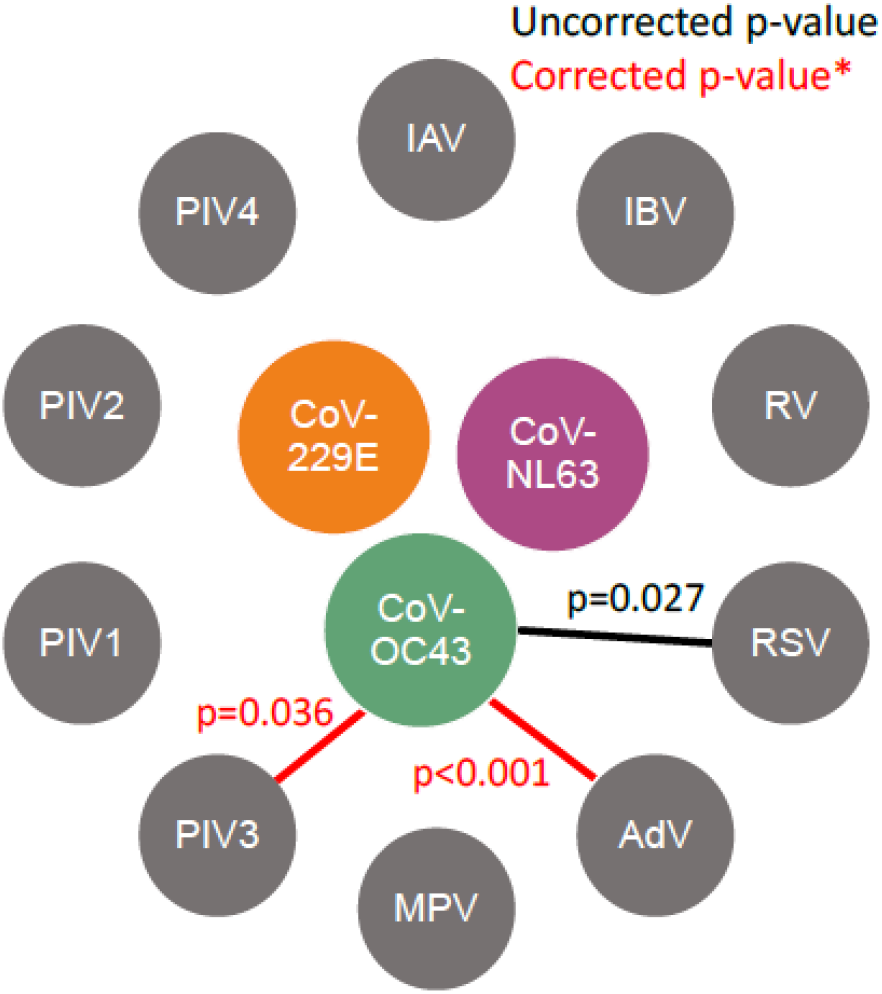
Interactions between human coronaviruses (CoV) and other respiratory viruses inferred using multivariable logistic regression based on 16,991 respiratory virus-positive patients. See supplementary Tables S8-S10 for full results. *p-values corrected for multiple comparisons using Holm’s method. CoV=human coronavirus; IAV=influenza A virus (H1N1 and H3N2); IBV=influenza B virus; RV=human rhinoviruses; RSV=respiratory syncytial virus; AdV=human adenoviruses; MPV=human metapneumovirus; PIV1=parainfluenza 1 virus; PIV2=parainfluenza 2 virus; PIV3=parainfluenza 3 virus; PIV4=parainfluenza 4 virus. Note PIV types were aggregated into PIV1/PIV3 and PIV2/PIV4 (genus groups) for the CoV-229E analysis, and PIV2 was excluded from the CoV-NL63 analysis.

In contrast, CoV-OC43 and CoV-NL63 generally exhibit annual periodicity of varying magnitude. A considerable degree of synchrony is observed in the timing of the peak prevalence of CoV-OC43 and CoV-NL63 for most seasons, whereas CoV-229E was more distinctive in its temporal pattern. For example, low levels of CoV-229E in 2007 coincided with high magnitudes of CoV-OC43 and CoV-NL63, whilst the largest prevalence of CoV-229E in 2010 coincided with low magnitudes of CoV-OC43 and CoV-NL63.

### Do coronaviruses interact with other respiratory viruses?

The cocirculation of sCoVs with other common respiratory raises the potential for ecological interactions, altering infection risks and the dynamics of population transmission. Our data did not permit analysis of potential within-host interactions among different sCoVs because of an absence of sCoV coinfections. We did, however, evaluate the potential for within-host interactions between sCoVs and other common respiratory viruses.

To do so, we analyzed the non-random mixing of respiratory viruses among virus-positive patients using multivariable logistic regression. We found a greater propensity for CoV-OC43 to coinfect with RSV (OR=1.68, 95% CI=1.05-2.63, uncorrected p=0.027), AdV (OR=2.93, 95% CI=1.87-4.5, uncorrected p<0.001), and PIV3 (OR=2.38, 95% CI=1.28-4.17, uncorrected p=0.004) (supplementary Table S8). The associations with AdV and PIV3 were supported following correction of p-values for multiple comparisons (p<0.001 and p=0.036 respectively).

No evidence of interactions with other respiratory viruses was found for either CoV-229E or CoV-NL63. Assessment of parainfluenza virus (PIV) types was limited by small numbers of coinfections; these viruses were aggregated at the genus level for the CoV-229E analysis, and PIV2 was excluded from the CoV-NL63 analysis. See supplementary Tables S9-S10 for details. The finding for PIVB must be treated with caution owing to 95% CI overlapping 1. Average age-specific predicted probabilities of sCoV coinfection for individuals with and without coinfection with each specific respiratory virus is given in Table 1.

**Table 1:** Age-specific predicted probabilities of CoV-OC43 detection among patients with and without putative interacting viruses

| Interacting virus | PCR-positive | Age group | Predicted probability* | Lower 95% CI† | Upper 95% CI† |
| --- | --- | --- | --- | --- | --- |
| AdV | No | <1y | 0.014 | 0.008 | 0.025 |
|  | Yes |  | 0.038 | 0.021 | 0.067 |
|  | No | 1-5y | 0.011 | 0.006 | 0.020 |
|  | Yes |  | 0.031 | 0.018 | 0.054 |
|  | No | 6-16y | 0.006 | 0.003 | 0.014 |
|  | Yes |  | 0.017 | 0.007 | 0.036 |
|  | No | 17-45y | 0.008 | 0.004 | 0.015 |
|  | Yes |  | 0.018 | 0.010 | 0.035 |
|  | No | 46-64y | 0.008 | 0.004 | 0.016 |
|  | Yes |  | 0.020 | 0.010 | 0.042 |
|  | No | >64y | 0.012 | 0.007 | 0.022 |
|  | Yes |  | 0.030 | 0.017 | 0.054 |
| PIV3 | No | <1y | 0.017 | 0.010 | 0.029 |
|  | Yes |  | 0.021 | 0.011 | 0.041 |
|  | No | 1-5y | 0.015 | 0.009 | 0.026 |
|  | Yes |  | 0.020 | 0.010 | 0.039 |
|  | No | 6-16y | 0.007 | 0.003 | 0.017 |
|  | Yes |  | 0.008 | 0.003 | 0.019 |
|  | No | 17-45y | 0.008 | 0.004 | 0.016 |
|  | Yes |  | 0.010 | 0.005 | 0.021 |
|  | No | 46-64y | 0.008 | 0.004 | 0.016 |
|  | Yes |  | 0.009 | 0.004 | 0.019 |
|  | No | >64y | 0.012 | 0.007 | 0.022 |
|  | Yes |  | 0.013 | 0.007 | 0.026 |

\* Predicted probability generated from fitted multivariable logistic regression
models based on 16,991 respiratory virus-positive patients. See supplementary
Tables S8-S10 for full analysis results. †CI=confidence interval.

## Discussion: Implications for COVID-19

The likely long-term impact of the recently emerged COVID-19 is a topic currently shrouded in uncertainty for countries worldwide. At the time of writing, global cases are mounting, with evidence of community transmission for a growing number of countries. In the absence of setting-specific data, an epidemiological understanding of related and unrelated co-circulating pathogens is prudent to guide preliminary estimates of who is at risk and when, and to develop research priorities pertaining to population susceptibility. Epidemiological knowledge of seasonal human coronaviruses (sCoVs) is lacking for many settings due to an absence of inclusion in routine diagnostic testing. Here, we described several key features of sCoVs based on a unique dataset derived from multiplex PCR diagnostic testing of a large, well-defined population over a thirteen year period.

It is well known that sCoVs cocirculate endemically with other common respiratory viruses, and coinfections are frequently observed [11, 12, 16]. In the West of Scotland, sCoVs typically peak in winter months alongside influenza viruses and RSV, as previously described [12]. The common occurrence of sCoVs during periods of high influenza activity highlights the importance of considering these viruses in the differential diagnosis of viral respiratory infections. This is particularly pertinent in the context of COVID-19 emergence; cocirculating viruses are associated with a broad spectrum of clinical presentation overlapping that of COVID-19, raising the potential for a large number of undiagnosed or misclassified cases in settings lacking the capacity for multiplexed testing. Currently in NHS Greater Glasgow and Clyde, Scotland, the majority of COVID-19 testing is being conducted in the hospital setting, where all cases of respiratory illness are simultaneously tested for influenza, RSV, adenovirus and *mycoplasma pneumoniae* using a multiplex panel, and severely ill patients receive a full multiplex that includes sCoVs.

Sex-specific numbers of sCoV differed by healthcare setting; we observed a trend towards female sCoV cases for primary care, but male sCoV cases for secondary/tertiary care. This finding may reflect sex differences in healthcare seeking behaviors and/or illness severity. In terms of sex differences in detection odds (a proxy for infection risk), a statistically significant sex effect was found for CoV-229E, with a greater odds among males than females. This finding is consistent with previous reports of a male bias for CoV-OC43 and CoV-NL63 among the hospital setting [12], and in relation to influenza hospitalizations and mortality in the context of acute respiratory infections more generally [22, 23]. It has been proposed that differences in sex hormones may explain variation in respiratory infection susceptibility, with testosterone exerting an immunosuppressive effect in males, and with estrogen playing a protective role in females [24, 25]. We note however our analyses do not control for potential confounding factors such as smoking or chronic obstructive pulmonary disease (COPD).

A greater number of sCoV cases were observed in children under five years of age and the elderly, particularly among patients admitted to hospital. This pattern is consistent with overall testing trends, and may therefore reflect the healthcare seeking behavior of concerned parents, clinician testing practices, and/or the burden of other respiratory agents, rather than a greater risk of infection. However, it should be borne in mind that these analyses were based on a patient population; the true community burden of mild and/or asymptomatic infections is likely to be much higher [26]. When aggregated into epidemiological groupings, age-specific probabilities of virus detection varied across sCoV types.

In contrast to that observed for sCoVs, there are relatively few COVID-19 cases thus far in children [27]. In the context of our study population, COVID-19 is closely related to CoV-OC43 (both betacoronaviruses), the most prevalent sCoV detection among patients under five. It is possible that pre-existing cross-immunity confers protection and/or attenuates the severity of COVID-19 leading to fewer tested/hospitalized children. The comparatively lower proportion positive and detection odds for school-aged children compared to younger children may potentially reflect a sustained level of CoV-OC43 immune-mediated protection, whereas waned immunity is expected to leave adults more vulnerable to coronavirus infections [27]. Assuming some degree of cross-immunity with COVID-19, our data are consistent with an expected fewer cases of COVID-19 in children but more cases among the adult population. Immunosenescence may exacerbate low levels of protective immunity in the elderly [29, 30].

Three key features of our data seemingly support the proposition of cross-immunity. Firstly, the contrasting age patterns of detection probabilities between closely related CoV-229E and CoV-NL63 (both alphacoronaviruses) may reflect niche segregation. Secondly, closely related CoV-229E and CoV-NL63 also displayed asynchronous seasonality, in contrast to CoV-OC43 and CoV-NL63, supporting a competition dynamic. Although our data did not permit in depth analysis of CoV-HKU1, others have reported differences in the timing of peak detections with CoV-OC43 (both betacoronaviruses) [12]. Thirdly, coinfections among sCoVs were not recorded in this study population, although detected by others albeit at a very low frequency [12]. More work is needed to establish whether low coinfection frequency among sCoVs supports an immune-mediated competition for hosts, or whether this reflects a limitation of diagnostic data which only captures a snapshot of an individual’s infection.

To our knowledge, evidence of immunological cross-protection between human coronaviruses is lacking, and reports of antigenic cross-reactivity are inconsistent. The potential for serological cross-reactivity between SARS-CoV and sCoVs has been shown by some [28, 29] but not others [30]. And although confinement of cross-reactivity at the coronavirus genus level is possible, consistent with the greater genetic relatedness of these viruses [31, 32], more general cross-reactivity between CoV-OC43 and CoV-229E has also been found [33]. Population serological surveys will be critical for establishing the true burden and age distribution of sCoV infections in the community and the potential for cross-protective immunity.

It should be borne in mind that the implications of population levels of cross-immunity are likely to vary according to the local epidemiological context. We note the predominance of CoV-OC43 detections previously observed in a comparatively urban but different Scottish patient population [14], a pattern more generally consistent at the Scottish national level [27]. Other trends observed within our study population are consistent with the Scottish national level, such as peak levels of CoV-229E detection in 2010 coinciding with low levels of CoV-NL63 and CoV-OC43 [27]. A predominance of CoV-OC43 has also been observed in Sweden [16], suggesting a potential consistency in sCoV dominance over wider geographical areas. However, differences are also apparent; for example, the relatively common detection of CoV-HKU1 in a different Scottish population [14] in contrast to ours, as also suggested in a recent report on respiratory virus detections in France [28].

Our study also highlights the potential for interactions between sCoVs and other respiratory viruses. In previous extensive analyses of virus-virus interactions, we found a strong signal of positive interactions at the within-host scale between human coronaviruses overall and respiratory syncytial virus (RSV), adenovirus (AdV) and parainfluenza viruses (combining types 1 and 3; human respirovirus genus) [6]. Here, our more in-depth analysis corroborates positive interactions at the sCoV type level. For CoV-OC43, the most prevalent in this study population, our results support our earlier findings of a higher propensity of coinfection with RSV, AdV and PIV3 than with other respiratory viruses. Note, these analyses were based on routine diagnostic testing and therefore the directionality of effects could not be determined. The association with RSV was not supported following a correction for multiple comparisons, although previous studies also report a high proportion of sCoV coinfections with RSV [12, 34]. The role of viral coinfections in the severity of acute respiratory illness remains a controversial topic [28–32]. Further work is needed to establish the role of viral coinfections in COVID-19 severity.

## Conclusions

The public health impact of COVID-19 is likely to vary according to the epidemiological context and healthcare infrastructure of the population. Our findings suggest continued monitoring of cocirculating respiratory viruses will be important for guiding accurate case ascertainment, research priorities surrounding population susceptibility, and for assessing the comparative population and healthcare burden of COVID-19 in the context of multiple cocirculating respiratory pathogens. Further work is needed to identify the mechanism of interactions between human coronaviruses and other respiratory viruses, and the role of viral coinfections in COVID-19 severity and burden.

## Data Availability

The patient-level data used in this study are available upon request to NHS Scotland (https://www.informationgovernance.scot.nhs.uk/pbpphsc/home/for-applicants/). Aggregated forms of summary data may be made available upon request to the corresponding author.

## Acknowledgments

This work was funded by the Medical Research Council of the UK (Grant MC_UU_12014/9). We are grateful to Louise Matthews, Richard Reeve and Paul Johnson for previous helpful discussions on inferring virus-virus interactions from patient-based data, and we thank Beatrix von Wissmann for her critique of an earlier draft of the manuscript. Author contributions: SN: conceptualization, data curation, methodology, investigation, formal analysis, visualization, writing – original draft; AH: investigation, writing – review & editing; DFPM: investigation, resources, writing – review & editing; JM: resources, writing – review & editing; RNG: resources, writing – review & editing; PRM: funding acquisition, investigation, writing – review & editing.

